# COVID-19 and Mental Health in China: Effects of Personality and Demographics

**DOI:** 10.1101/2023.01.20.23284600

**Authors:** Xiao Zhang, Michele Battisti, Eugenio Proto

**Author notes:** Corresponding author,uk.

## Abstract

China was the first country affected by the COVID-19 virus and it reacted strongly in the first months of 2020. This paper presents new evidence on the deterioration in mental health in China between 2018 and 2020. Using two waves of the China Family Panel Studies (CFPS) we can follow the same individuals pre and during the pandemic periods. We find clear evidence of a moderate level of mental health deterioration between 2018 and 2020. The prevalence of severe cases of depression, measured using an eight-item version of the common CES-D scale, increased from 6.33% in 2018 to 7.54% in 2020; quantifiable as around a 19% increase. This deterioration is higher for individuals who are subject to strict lockdowns, about 0.3 symptoms more on average, and it is stronger among those who already reported symptoms of depression in the 2018 wave of data. The effects we find are larger for individuals with more open personalities: one standard deviation of the Openness trait corresponds to 0.08 more symptoms, while more Neurotic individuals do not seem to be more affected. Younger cohorts and individuals with lower levels of education are more affected. Males seem slightly more affected than females, although this difference is statistically non-significant.

## Introduction

China, when compared to most Western countries, has been particularly effective in controlling the spread of COVID-19 over the last three years^2^. However, the effects of the pandemic on mental health, particularly regarding the stringent procedures put in place to control COVID-19, has received far less attention. More generally, a recent *Lancet* editorial emphasizes how mental health in China is a particularly compelling issue “*because it plays out against the wider backdrop of mental health disorders in China, which remain largely unaddressed*” [1].

Therefore, given the size of the country and the potential extent of the problem, it is crucial to identify the individuals that may have been more affected in terms of mental health for several reasons. Firstly, this can lead to identification of at-risk groups, as well as more personalized psychological or psychiatric treatments, even for the post-COVID period. Secondly, understanding how different individuals with different personalities have reacted in China, and comparing this reaction with individuals in Europe and the United States, may improve our understanding of how mental health is affected by extreme situations and the link between personality and mental health.

We analyze mental health deterioration in China during the first part of the pandemic, using two waves of the China Family Panel Studies (CFPS). The CFPS covers a representative sample of the entire Chinese population. The first CFPS wave consists in data collected in 2018 (i.e., before the pandemic), while the other wave of data was collected from July to December 2020 (i.e., the pandemic wave). The panel structure of the CFPS dataset allows us to compare the same individuals over time, i.e., before and during the pandemic. Our analysis shows clear evidence of a significant increase in the prevalence of serious cases of depression between 2018 and 2020.

The CFPS dataset includes a set of standard personality questions derived from the most widely used taxonomy of personality, the “Big Five” model as measured by the Revised NEO Personality Inventory [2]. Thanks to this information, we were able to test how personality interacts with the pandemic to affect mental health. We find that the worsening of depression symptoms is more common for individuals with a more open personality. On the other hand, neuroticism does not lead to a more serious mental health deterioration, as one may naturally expect (see [3] for a thorough description of the Big Five personality traits). As we discuss below, this is remarkably in line with the previous analysis on mental health effect of the Covid-19 pandemic, based on very different contexts and environments. This invariance lends more support to the notion that the Big Five personality traits are a universally common feature in all human being, in addition it can provide further insights into the large literature analyzing the link between personality and mental health.^3^

Concerning basic demographics, younger cohorts are more affected than older ones. Individuals with lower education also report more mental health deterioration compared to individuals who report higher levels of education. We do not observe any significant difference between genders. We will discuss in the final section of this article how this compare with the other existing analysis of the effect of the COVID pandemic on mental health.

Moreover, using information on the province of residence at the time of the pandemic, we find that individuals that have been subject to strict lockdowns report a higher level of mental health deterioration; the observed deterioration during lookdown is largely due to changes among individuals who reported some depression symptoms before the pandemic, rather than among individuals who did not report any symptom in the pre-pandemic wave from 2018.

There are very few country-representative studies on the effect of the COVID pandemic on wellbeing in China. To the best of our knowledge only two: [6] perform a Chinese country-representative study that compares emotional wellbeing from before the pandemic to the pandemic period. Their main goal is to analyze emotional wellbeing (or Happiness), rather than a specific measure of mental health (such as depression or anxiety). They find that the beginning of the coronavirus epidemic led to a 74% drop in overall emotional well-being. The other is [7] that, like the current study, is based on the CFPS dataset. They examine the effect of COVID incidence on the mental health of individuals in different provinces (whereas we focus on the difference in average between the pre-COVID period and the first COVID wave). Their results are fully consistent with ours, although [7] do not analyze how personality affects mental health deterioration, as we do.

Several studies find some mental health deterioration in China among specific groups during the pandemic. These contributions focus on specific groups rather than on the whole population: COVID-19 patients ([8] [9] [10]); university and college students ([11] [12] [13] [14]); children and adolescents ([15] [16] [17] [18]); or other subgroups of the population ([19] [20] [21] [22] [23]). Furthermore, several of these China-based contributions use data collected during the pandemic through non-representative and mostly online surveys (e.g. [24] [25] [26] [27]). Finally, there is a large and growing literature investigating the effects of the pandemic on mental health using data from several countries ([28] [29] [30] [31] [32] [33] [34] [35] [36] [37] [38] [39]).

We well explain how some of the above-mentioned contributions relate to our work in the final Discussion section.

## Results

The left-hand panel of Figure 1 below shows a significant increase in the prevalence of serious depression symptoms between the pre-pandemic (2018) and the first pandemic year (2020). We will refer to this difference as mental health deterioration henceforth. The prevalence of severe case of depression over our weighted sample is 6.33% (95% CI 5.98, 6.68) in 2018 and 7.54% (95% CI 7.16, 7.93) in 2020.

**Figure 1.**
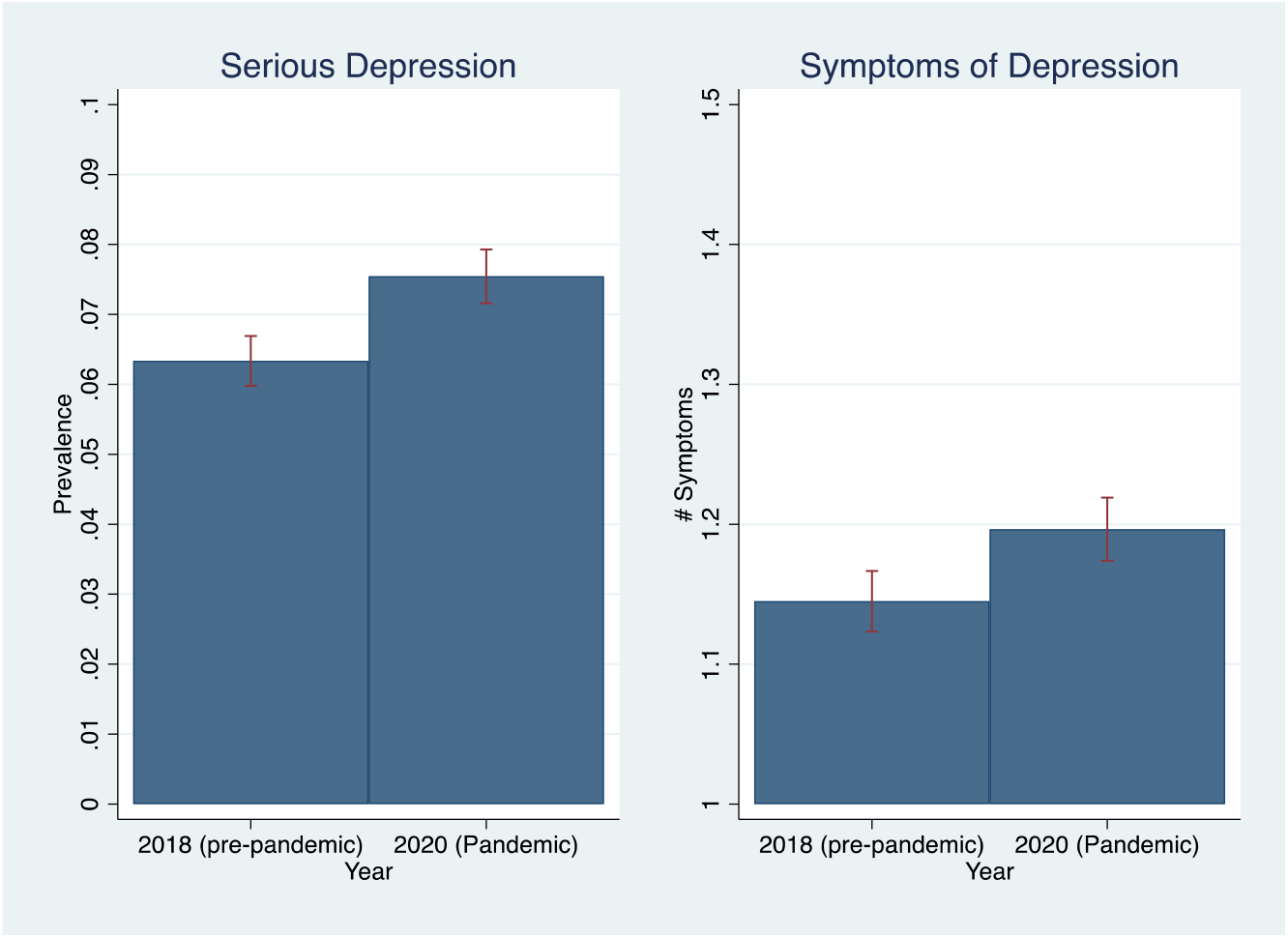
Depression pre and during the pandemic years: In the left panel, serious depression is defined when responders report 12 or more symptoms in the CES-D questionnaire. The right panel reports the average number of symptoms reported. The bars represent the 95% confidence intervals. Statistics are weighted using the survey sample weights. Data source: CFPS dataset, 2018 and 2020 waves. All statistics are produced using a weighted sample.

The right-hand panel of Figure 1 investigates differences in the average symptoms of depression, which are measured using the eight-item CES-D scale, rising from 1.144 (65% CI 1.123, 1.166) in 2018 to 1.196 (95% CI 1.174. 1.219) in 2020, as shown on the right-hand panel of Figure 1. This implies a difference of 0.051 (95% CI 0.016, 0.087), which is significantly different from zero (*p-value* = 0.004).

One possible concern is that differences between 2018 and 2020 are simply driven by the fact that respondents are two years older in 2020 since age might affect the prevalence of severe depression. We address this concern by estimating the effect of the COVID pandemic on our mental health indicators (serious depression and symptoms) by controlling for Age and Age Squared. The result is presented in Figure 2. From the top panel we note that the difference of effect before and after controlling for age is quite small (from 1.2% to 0.9%). From the bottom panel, we note a more substantial difference in terms of symptoms (from 0.051 to 0.028) that becomes non-significant. Therefore, these results suggest that age affects the reporting of symptoms more in the extensive margin (i.e., responders with symptoms report more of them) than in the intensive margin (i.e., responders report symptoms for the first time), while the opposite is true in the COVID period. Hence, responders who did not report symptoms previously begin to report them significantly more in the COVID period, independently from their ages.

**Figure 2:**
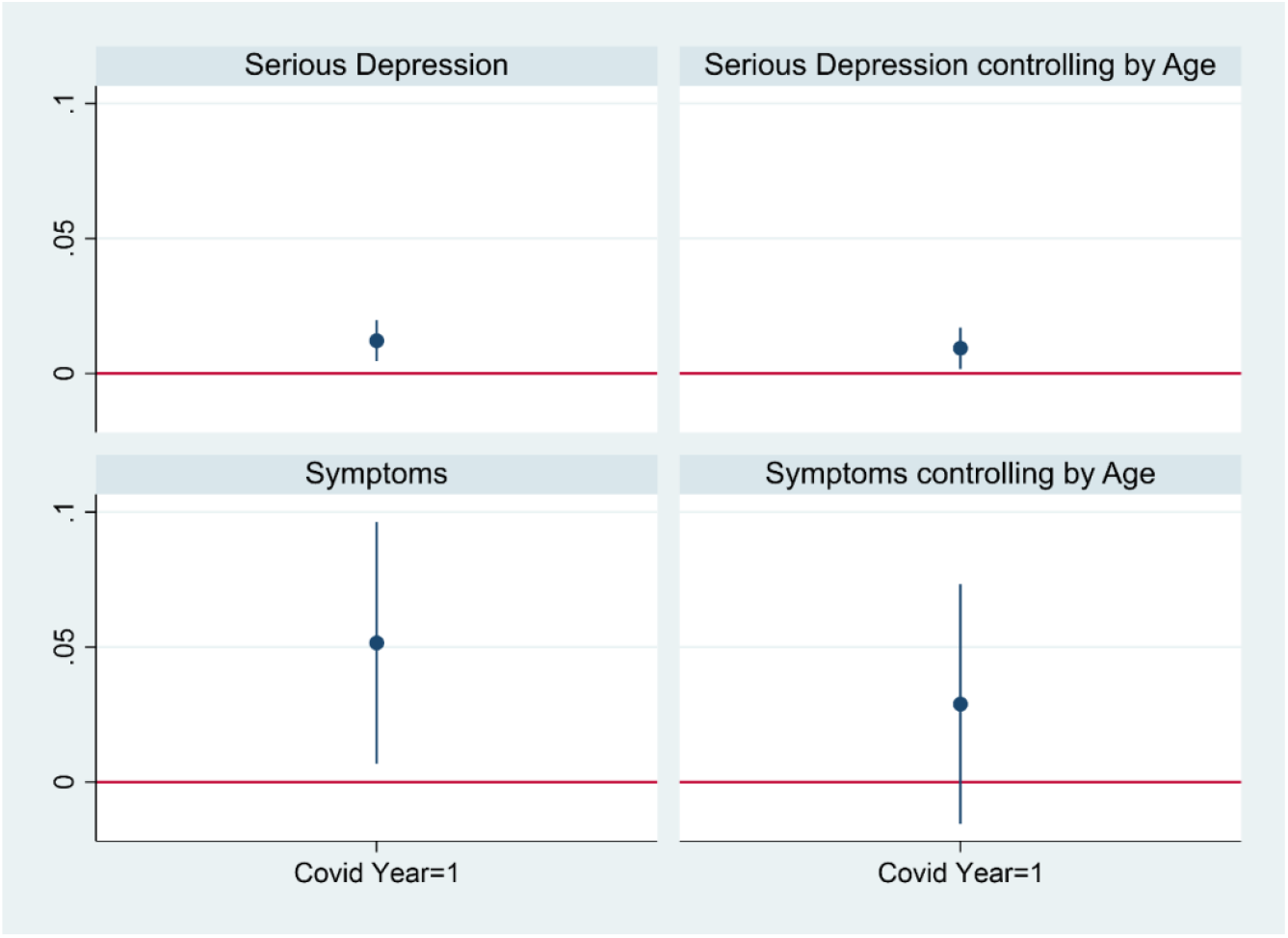
Estimated differences in the depression pre and during the pandemic controlling by Age. The table reports the OLS coefficients of the COVID wave dummy in 4 separate regressions. In the top panels, the dependent variable is the case of serious depression. In the bottom panels, the dependent variable is the symptoms reported. The regressions in the right panels include the variables Age and Age squared (unreported in the table), while the regressions in the left panels do not. The bars represent the 95% confidence intervals with robust standard errors. The 4 regressions include the same responders, and the data sample is weighted to generate population representative estimation.

### Mental health deterioration and personality traits

Next, we will analyze whether the mental health deterioration we have outlined above has been experienced more acutely by individuals according to their different personality traits, by estimating a multiple linear regression model (further illustrated in the materials and methods section).

Our results are presented graphically in Figure 3 below, demonstrating the regression’s coefficients with and without additional regressors to control for potential confounding variables (the full regressions are presented in Table S4 of Supplementary Information). From both regressions we note a strong effect of Openness (see left panel of Figure 3, corresponding to Column 1 of Table S4 of Supplementary Information), where one standard deviation of Openness corresponds to 0.08 more symptoms. This magnitude is similar to the one reported in the right panel of Figure 3 (corresponding to Column 3 of Table S4 of Supplementary Information), where we add control for several demographic factors. Neuroticism is the other significant coefficient, negative and equal to -0.5, which may look surprising, but it is roughly in line with other studies analyzing how different personalities reacted to the pandemic, as argued in [33] and in the final section of this article.

**Figure 3:**
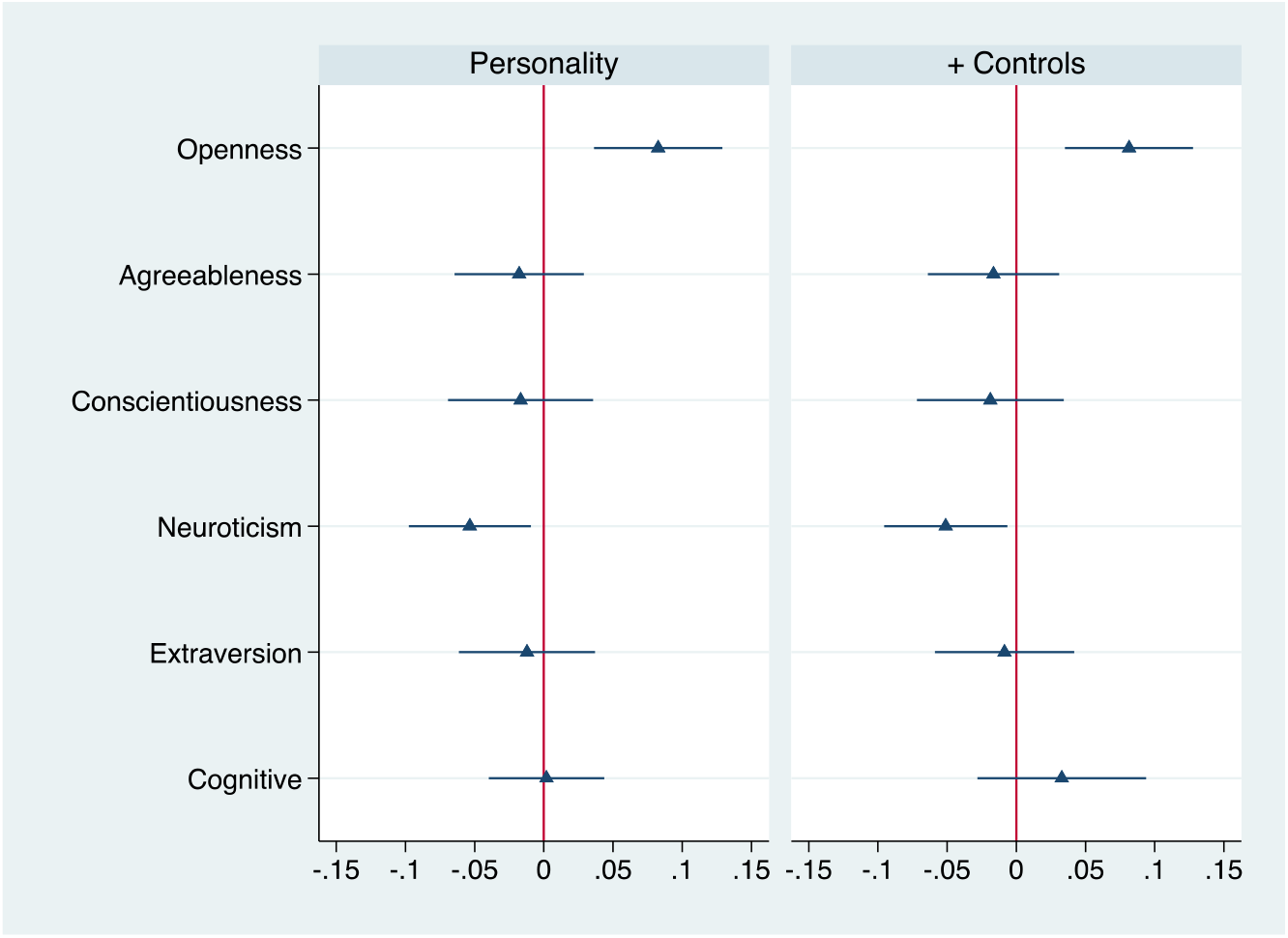
Mental health deterioration with respect to different personalities. The table reports the OLS coefficients of the two regressions where the dependent variable is the difference between the number of symptoms in 2018 and 2020. The personality trait measures are standardized. The bars represent the 95% confidence intervals with robust standard errors. Data are weighted to generate population representative estimation. Gender, age group, urban residence, family size, and months of the interview are included in the regression related to the right panel but omitted in the figure. The complete estimation of the two regressions is presented in Table S4 of the Supplementary Information.

### Mental health deterioration by basic demographic variables

Figure 4 investigates the same statistics described in Figure 1 but distinguishes between men and women. The left-hand panel of Figure 4 shows a significant increase in the prevalence of serious depression among women, rising from 7.50% (95% CI 0.069, 0.080) in 2018 to 8.41% (95% CI 0.078, 0.090) in 2020. Among men, the prevalence of serious depression increases from 0.052 (95% CI 0.047, 0.057) in 2018 to 0.067 (95% CI 0.062, 0.072) in 2020. Both differences are statistically significant at the 5% level, with *p-value*<0.00 for men, and *p-value* = 0.024 for women. This is mirrored in the right-hand panel, where we observe an increase in symptoms of depression for both genders.

**Figure 4.**
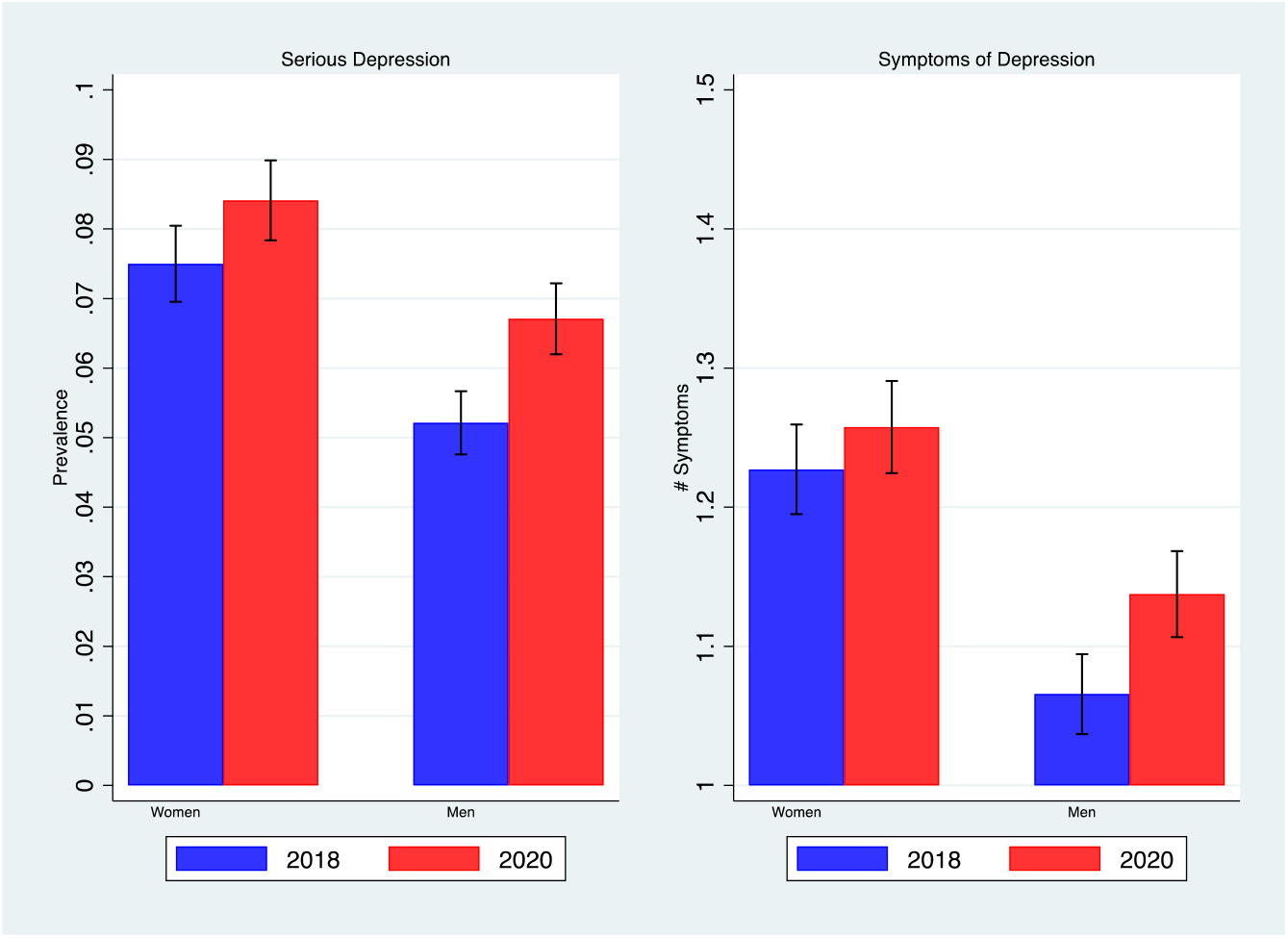
Depression pre and during the pandemic years by gender: In the left panel, serious depression is defined when responders report 12 or more symptoms in the CES-D questionnaire. The right panel reports the average number of symptoms reported. The bars represent the 95% confidence intervals. Statistics are weighted using the survey sample weights.

Figure 5 investigates the prevalence of serious depression as well as depression symptoms for different age groups in 2018 and 2020. The left panel shows an increase in serious depression prevalence in all age groups. Quantitatively, the increase is larger among younger cohorts. Among individuals younger than 18 years of age, the prevalence of serious depression rises from 1.52% (95% CI 0.009, 0.020) in 2018 to 3.98% (95% CI 0.030, 0.049) in 2020, while among the oldest cohort (age 65 and above), it rises from 9.44% (95% CI 0.081, 0.107) in 2018 to 10.37% (95% CI 0.092, 0.115) in 2020. From the right panel of Figure 4, we note that symptoms of depression have increased in all age groups, except for those aged 40-65.

**Figure 5.**
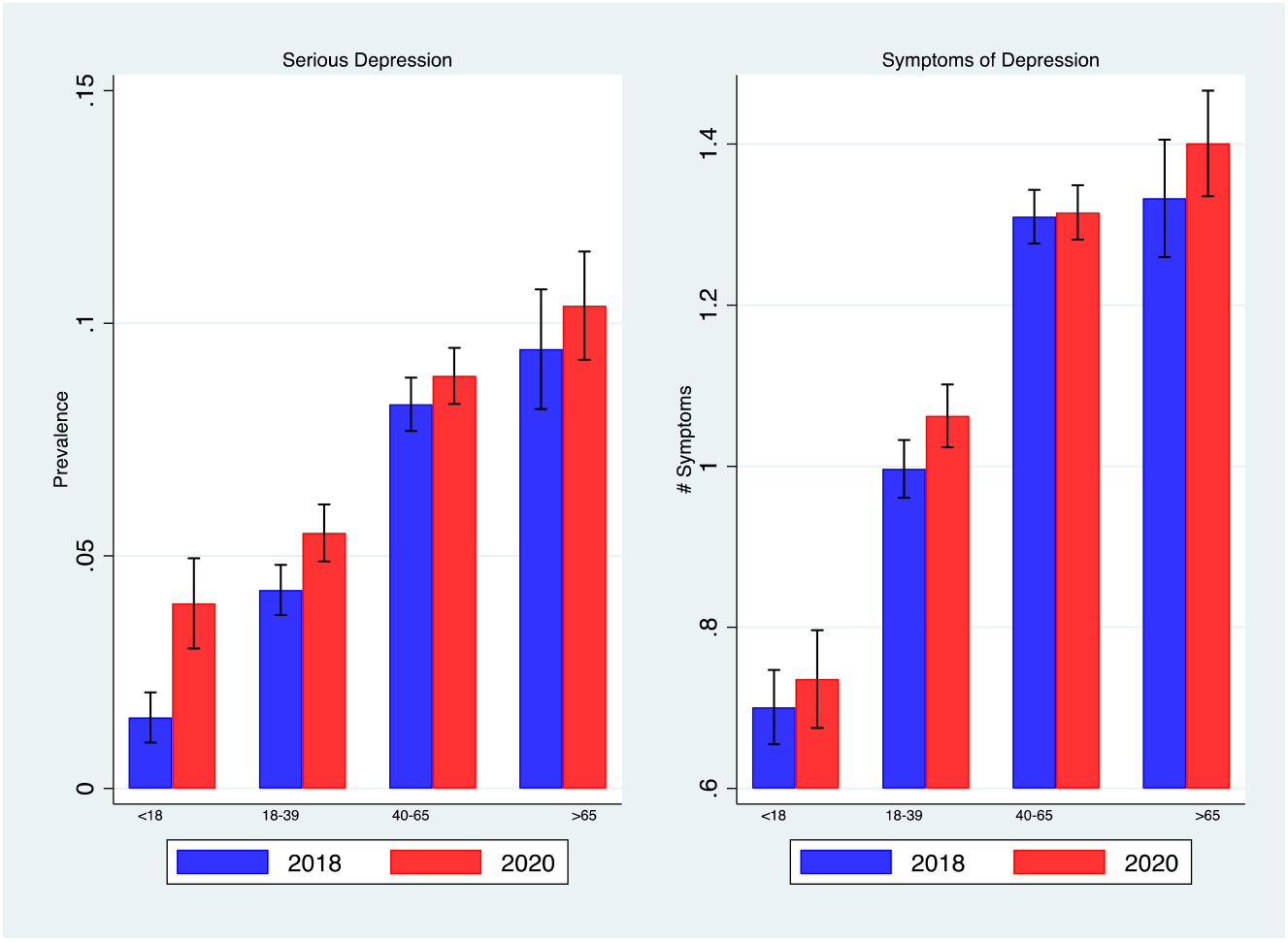
Depression pre and during the pandemic years by age groups: In the left panel, serious depression is defined when responders report 12 or more symptoms in the CES-D questionnaire. The right panel shows the average number of symptoms reported. The bars represent the 95% confidence intervals. Statistics are weighted using the survey sample weights.

Figure 6 investigates the differences in the levels of education. The left panel shows that there is an increase in the prevalence of depression for individuals with a primary school degree: from 8.90% (95% CI 0.082, 0.095) in 2018 to 11.28% (95% CI 0.106, 0.120) in 2020, and a slightly smaller rise for those with a secondary school degree: from 5.09% (95% CI 0.046, 0.056) in 2018 to 6.02% (95% CI 0.055, 0.065) in 2020. Depression is relatively stable among those with tertiary education at around 4%. The right panel features the same decreasing patterns in terms of symptoms differences between the two waves, showing larger differences among those with lower levels of education.

**Figure 6.**
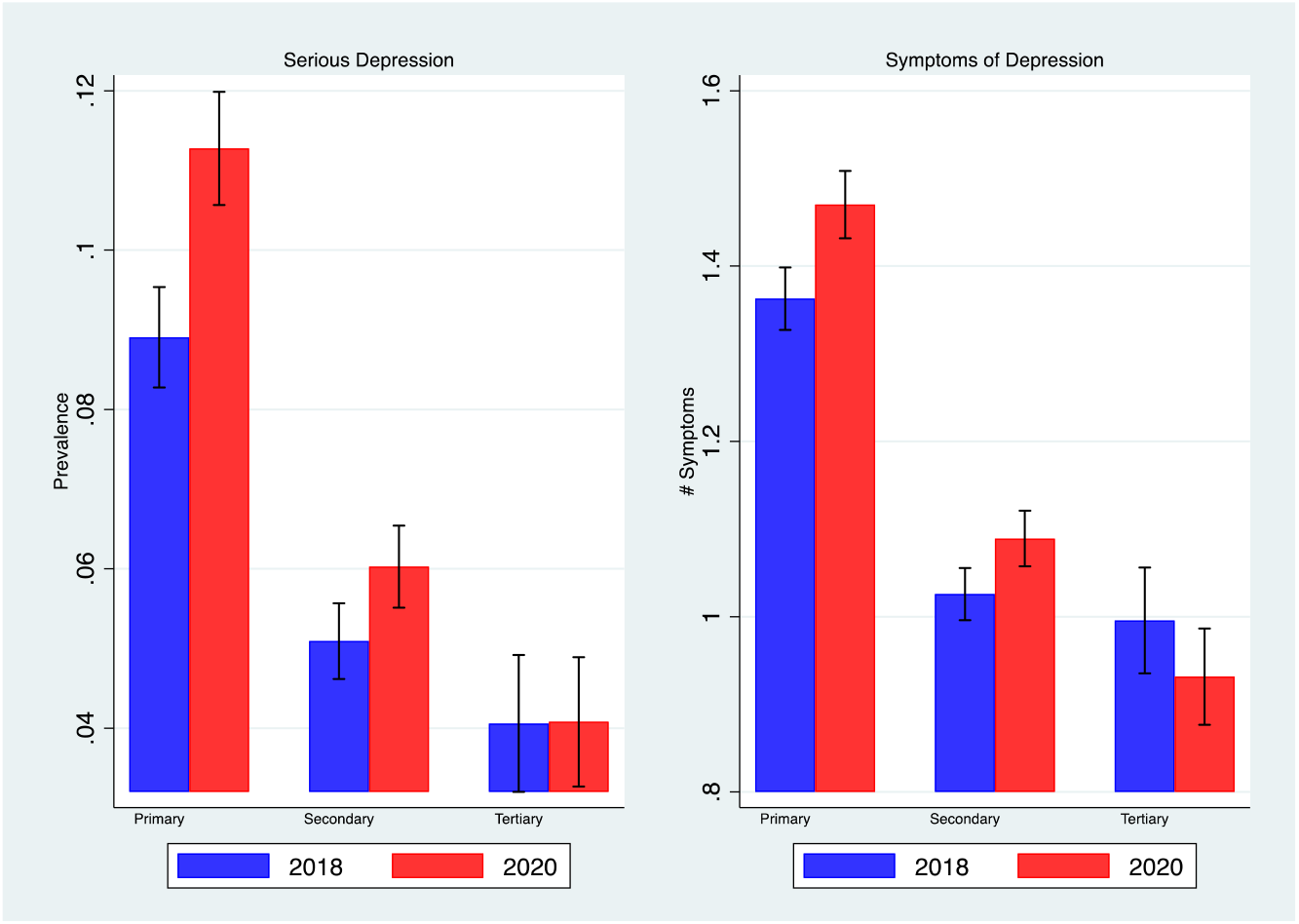
Depression pre and during the pandemic years by classes of education: In the left panel, serious depression is defined when responders report 12 or more symptoms in the CES-D questionnaire. The right panel reports the average number of symptoms reported. The bars represent the 95% confidence intervals. Statistics are weighted using the survey sample weights.

We further analyze whether mental health deterioration has been experienced more acutely by some groups compared to others by estimating a multiple linear regression model, which will allow us to control for other potentially confounding factors. Figure 7 reports the estimated coefficients of the regression presented in our material and methods section and includes the 95% confidence intervals (see Table S5 in the Supplementary Information for more detail).

**Figure 7:**
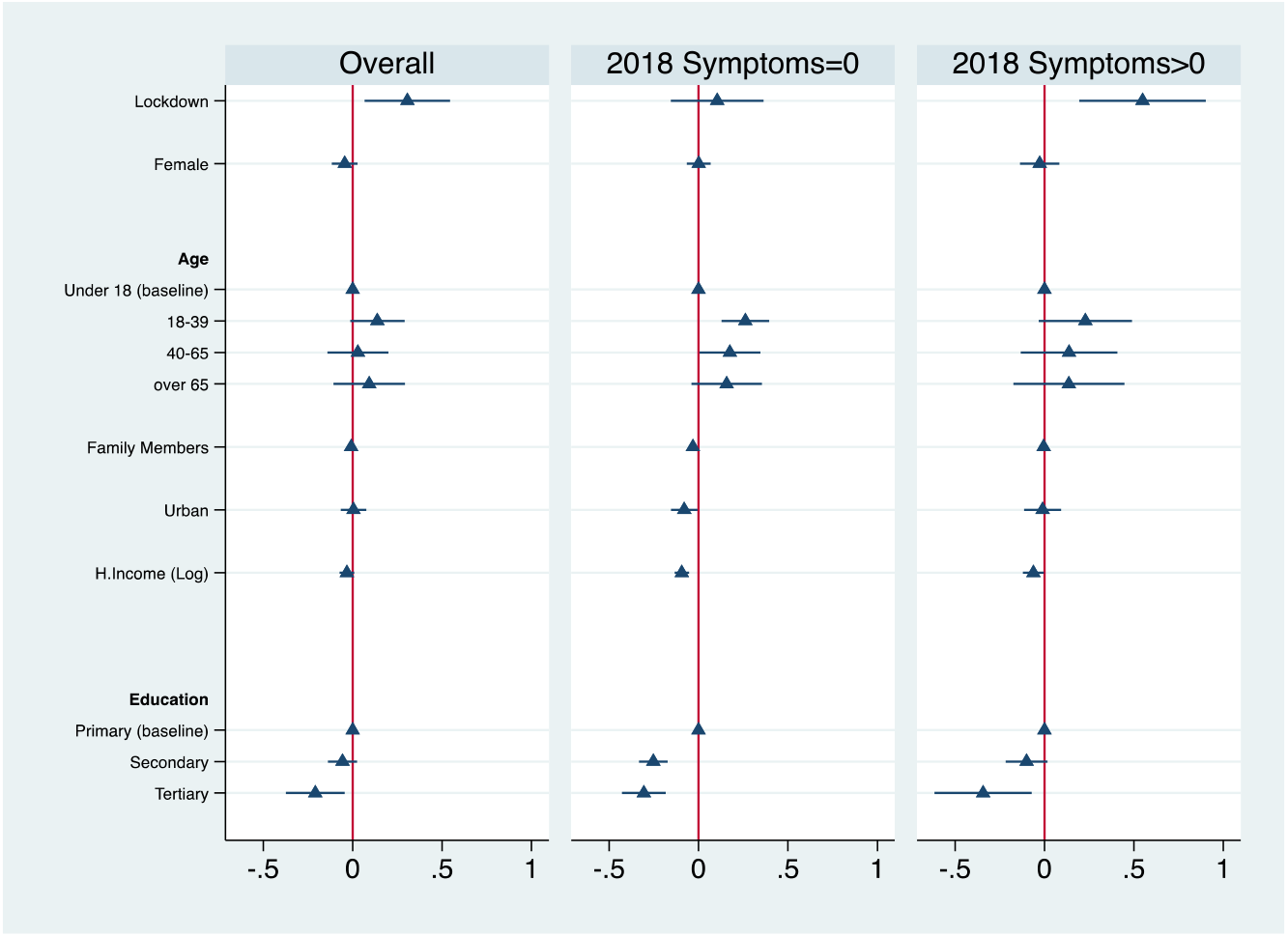
Mental Health Deterioration among different groups. The table reports the OLS coefficients of the three regressions where the dependent variable is the difference between number of symptoms in 2018 and 2020. The bars represent the 95% confidence intervals with robust standard errors. Data is weighted to generate population representative estimation. Marriage status and months of the interview are included in the regression but omitted in the figure. The complete estimation of the three regressions is presented in Table S5 of the Supplementary Information.

First, from the left panel, we note that individuals under lockdown measures (i.e., those living in the provinces of Hubei and Xinjiang) reported over 0.3 more symptoms (hence more than 1 in 4 individuals), compared to the central and the right panels. By comparing the central and the right panel of Figure 4, we note that the more affected individuals are the ones that were already reporting symptoms in the 2018 wave.

Figure 4 shows that Female responders seem to report less mental health deterioration than males. However, from Figure 7 (and Table S5 in the Supplementary Information), we note that this difference is not statistically significant (*p-value*= 0.230). Younger adults aged between 18 and 39 constitute the most affected age group. In particular, the central panel of Figure 7 shows that mental health deterioration tends to decline with age among subject over 18, especially among those that did not report any symptoms in 2018. Individuals with tertiary education reported significantly less deterioration than individuals with only primary education (more than 0.2 symptoms), and that deterioration increased among those with lower education levels (something we already noted above when analyzing Figure 6).

Finally, Column 4 of Table S5 of Supplementary Information presents differences with respect to Job Status: only individuals employed in the agrarian sector reported significantly smaller increases in symptoms than the baseline (i.e., those employed in the industry and in the other non-agricultural sectors). Column 5 of the same table in the Supplementary Information outlines no significant differences arising from provincial GDP per capita, provincial public expenditure or provincial population density.

## Materials and methods

### Data

As previously mentioned, we used the 2018 and the 2020 waves of the China Family Panel Studies (CFPS), which is a large panel dataset representative of the Chinese population. The 2018 wave had 37,354 total individual respondents, while the 2020 wave had 28,590. We used sampling weights as suggested by the data collectors. Given that they are available only for the 2018 wave we use the same when we analyze data from the 2020 wave. We use robust standard errors in all our regression analyses.

After removing individual data with missing mental health information on the CES-D questionnaire (9,160 in total), and those who were part of the survey in 2018 but not in 2020 for various reasons, we have 18,127 respondents appearing in both waves.

We will now discuss this attrition in more detail. Table 1 below reports the number of valid observations belonging to each wave and how many individuals are observed in both waves. Among individuals interviewed in 2018 with information on the CES-D and weights, 60.1% were re-interviewed in 2020. While an attrition rate of nearly 40% is substantial, it compares favorably with previous researches (e.g. [29][34]). In Table S3 of the Supplementary Information, we perform a simple attrition analysis which shows that using data from the 2020 wave results in a younger sample. This is to be expected since older respondents from 2018 are less likely to appear in the 2020 wave (the average age is 42.531, down from the initial 44.791 years).^4^ The average symptoms of depression slightly decrease in a consistent manner (the difference is non-significant at the 5% level). Average education and household income are also slightly higher, which is consistent with the age differences in this context.

**Table 1.** Cross-sectional and longitudinal dimensions.

| Wave | Individual is observed twice |  | Total |
| --- | --- | --- | --- |
|  | No | Yes |  |
| 2018 | 11,708 | 18,127 | 29,835 |
| 2020 | 6,398 | 18,127 | 24,525 |
| Total | 18,106 | 36,254 | 54,360 |

Source: CFPS (waves 2018 and 2020). The data are from China Family Panel Studies (CFPS), funded by Peking University and the National Natural Science Foundation of China. The CFPS is maintained by the Institute of Social Science Survey of Peking University.

Overall, we can say that the population represented in our final sample is slightly younger than a Chinese-representative sample.

There are two lockdowns imposed at the province level in China in 2020. The first one was imposed in Hubei province from 23^rd^ January to 8^th^ April 2020; while the second one was imposed in Xinjiang province in July and August 2020. Considering that the data in the 2020 wave of the CFPS was collected from July to December 2020, we distinguished two groups of responders who were affected by lockdowns (based on whether they have experienced lockdown before the interview or are during lockdown when they are interviewed): the first one is the responders in Hubei province for the entire 2020 wave (a total of 247 observations in the balanced panel), while the second one is the responders in Xinjiang province in July and August 2020 (for a total of 39 observations in the balanced panel). Due to the small sample size of the second lockdown, we have grouped both lockdowns together in our analysis.

In Table S1 and S2 of the Supplementary Information, we provide a summary description of all variables we use for each wave of the CFPS dataset that we will merge to perform our analysis.

### The 8-item Center for Epidemiologic Studies Depression Scale

To identify the levels and changes in mental health in the 2018 and 2020 waves, we used two measures based on the 8-item Center for Epidemiologic Studies Depression Scale, CES-D. The CES-D is a well-known instrument used for evaluating mental health where the respondent reports the extent to which each of the eight symptoms is present in the few weeks before the survey takes place. The respondent can pick from a few possibilities for each of these symptoms, based on weekly frequencies: rarely (less than 1 day), a little (one or two days), occasionally (three or four days), or often (five-seven days).^5^

Following common practice, we used a measure based on a binary indicator of being at risk of presenting with mental health problems (CES-D “caseness” score). In the 20-item questionnaire, the caseness is typically calculated after each answer is coded between 0 (no symptoms) to 3 (frequent symptoms) and then added up, so that the resulting score falls between 0 and 60. If the total score is 28 or above, the subject is considered to have severe depression. Accordingly, in the 8-item version – ranging from 0 to 24 – we assume that if individuals report a score of 12 and above (calculated considering the ratio 28/60 from the 20-item version) we classify them as having severe depression. In parts of our analysis, we also define a lower threshold for mild depression.

To measure the level of mental deterioration between the two waves, we counted the differences in symptoms between waves for each responder appearing in both waves, coding it a ‘symptom’ every time a specific way described in the questionnaire has been felt occasionally (three or four days), or often (five-seven days).

We assume that the total number of symptoms is a cardinal measure and therefore the level of deterioration (or the difference between the sum of symptoms in the pre-pandemic and pandemic waves) can be considered a cardinal measure as a simple linear OLS estimator can be used for the regression.

### Regression analysis

In the first part of the analysis, we compared weighted means and provided 95% confidence intervals. In the second part, we performed some regressions, using OLS to estimate different specifications of this linear econometric model:

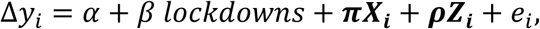

where ∆*y*_*i*_ is the change in the number of symptoms reported using the CES-D as defined above between 2018 and 2020 waves; *lockdowns* is a dummy where equals to 1 if the responder has experienced lockdown before the interview (in Hubei province for the entire 2020) or is interviewed during a lockdown (in Xinjiang province during July and August 2020); ***X***_***i***_ is a vector of demographic variables, including gender, age group indicators, month of interview indicators in 2018, household size in 2020, marital status in 2020, education indicators in 2018, and per capita household income in 2018. ***Z***_***i***_ is an employment status indicator in 2018. Finally, *e*_*i*_ is the error term of the regression. All regressions are estimated using 2018 weights and heteroscedasticity-robust standard errors.

## Discussion

There is widespread evidence that mental health has been severely affected by the COVID-19 pandemic ([28] [29] [43]), and it will likely be a major issue in the post-COVID world [44]. From the analysis presented in this article, we saw cases of serious depression in China increased by about 1.2 percentage points (about 19%). It is difficult to compare this with Europe and the US, where other scales have been used. To provide an idea of the possible differences, we calculated an index of mild depression defined as a number of symptoms larger or equal to 8 (as opposed to serious depression, where the threshold was 12). This demonstrated a significant increase from 25.2% to 26.9% as reported in Figure S1 of the Supplementary Information. We can compare this figure with [29], who find an increase of 13.5 percentage points in the UK from 24.3% in 2017-2019 to 37.8% in April 2020, based on GHQ-12 score ≥3. While the assessment method is based on different questionnaires, the increase in mental health problems in the UK seems to be much higher than in China, according to our data.

As noted above, it is crucial to identify at-risk groups and design personalized treatments, particularly in large countries such as China. People with more open personalities seem significantly worse-off, consistent with the view that certain personalities may be more likely to suffer the limiting of personal contacts and social life brought about by COVID-19. Openness is the trait that reflects preferences for exploration and new experiences (see e.g., [3]); in fact, this trait is often called “openness to experience”. Individuals with this type of personality seem to suffer more during a pandemic period, which is characterized by many constraints on individual freedom and restrictions on socializing with others at work and during leisure time. [33] found similar results in the UK.

On the other hand, neuroticism does not predict more deterioration in mental health. Neuroticism is linked to individuals’ higher sensitivity to negative outcomes and threats (see e.g. [3]) of the sort that have been pervasive in the current pandemic. As argued in [33], individuals with highly neurotic personalities have normally experienced several negative shocks during their lives; hence, there might be a sort of habituation effect at play. The effects of the pandemic on the mental health of different personalities are remarkably consistent with the rest of the literature on the subject, and we refer to [33] for a detailed discussion of this small but growing literature.

The Big Five traits-based personality questionnaire used in the CFPS is derived from the most widely used taxonomy of personality, as measured by the Revised NEO Personality Inventory [2]. The remarkably similar effects of the different traits on mental health during the pandemic across so different culture lends more support to the notion that they are invariant across European, North American, and East Asian samples, then suggesting their universality ([2][46]).

Considering the effect of pandemic across different ages, our finding that individuals who belong to the younger cohort are more affected compared with individuals in the older cohort is generally consistent with many country-representative studies. For the United States, [34] [35] [36] [37] report more severe deterioration of mental health among younger individuals compared with older individuals. For the UK [28] [29] [30] show evidence that younger individuals report more severe mental health deterioration compared with older individuals. [39] finds similar results in a study taking place in South Korea ^6^

With respect to the gender differences, our finding that males report only slightly and statistically insignificant more mental health problems than females is not conflicting with other studies based on the Chinese population [25] [49] [50]. This finding is also in line with [34] [35] using data from the United States, they do not report any significant difference between men and women. On the other hand, [36] [37] find that women are more affected by COVID-19 compared with men in the United States, and [28] [29] [31] [46] show that women’s mental health seem more affected by the pandemic in the UK. Overall, these mixed findings related to gender are in line with [48], who in an international meta-analysis did not find clear gender differences when comparing the mental health effects of the pandemic.

We find that individuals who have lower educational backgrounds and lower incomes are affected by COVID-19 more compared with individuals who have higher educational backgrounds and higher incomes^7^. However, the evidence on the differential effect on education are mixed in US-based studies. While [35] and [36] present evidence that the group with the highest increase rate in symptoms of anxiety and depression are the individuals with educational backgrounds lower than high school, [34] does not find a significant difference between individuals with and without college degrees, and [37] showed that individuals with secondary educational backgrounds reported the highest prevalence of depression during COVID-19. In addition, [37] find that individuals with lower income suffered more from COVID-19. Finally, UK-based studies typically find that the most significant mental health deterioration is among higher income and education groups ([28] [29] [31] [32] [46]).

These different findings on the effect of the pandemic among different genders and educational attainments in different countries would represent an interesting puzzle for further investigation.

## Data Availability

All data produced are available online at 
https://opendata.pku.edu.cn/dataverse/CFPS?language=en, upon request . 

## Acknowledgements

This paper uses data from China Family Panel Studies (Wave 2018 and Wave 2020). The CFPS was launched in 2010 by the Institute of Social Science Survey of Peking University. We thank co-authors and colleagues for their helpful comments and suggestions. Any errors in this article are the sole responsibility of its authors.

## Author contributions

Conceptualization: MB, EP

Data analysis: MB, EP, XZ

Writing - original draft: MB, EP, XZ

Writing - review and editing: MB, EP, XZ

## Supplementary Information

**Table S1:**
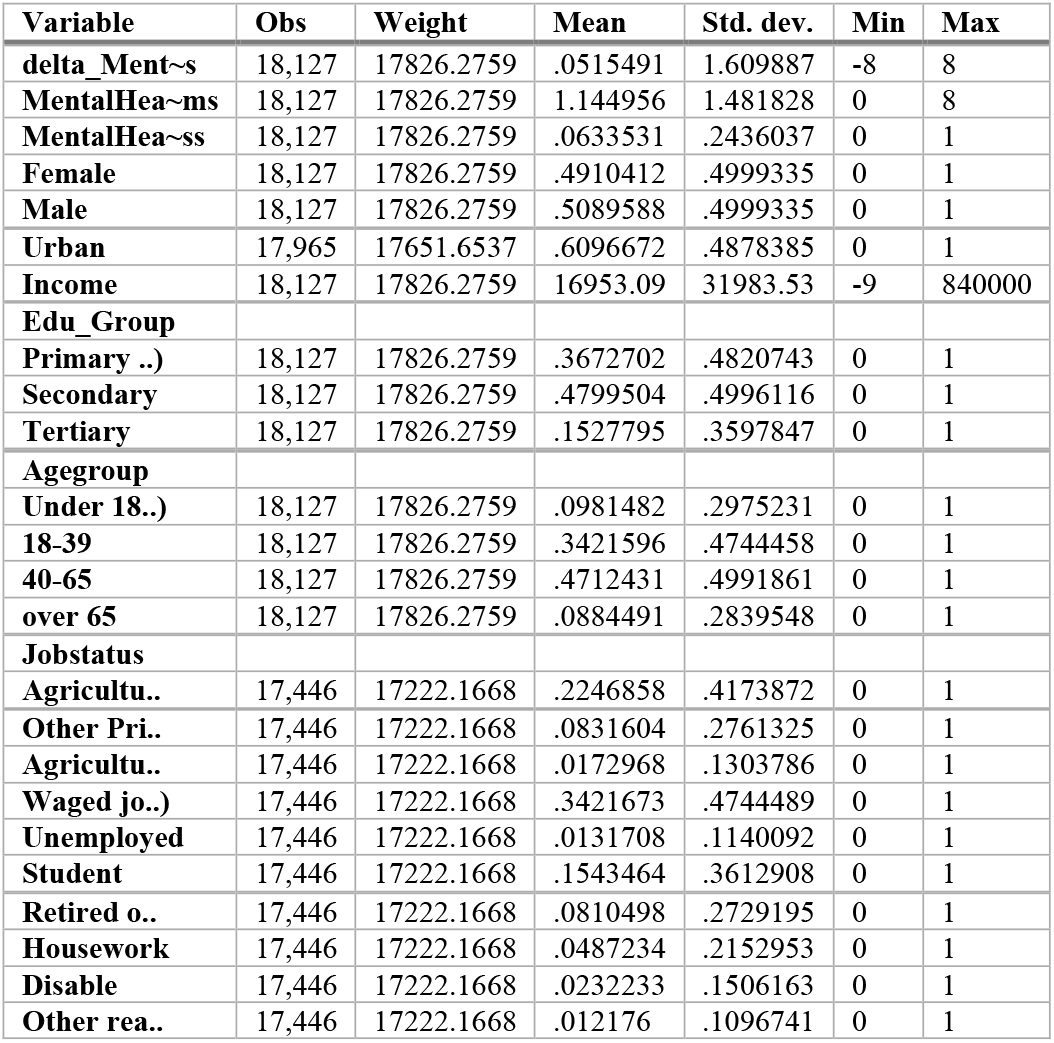
Descriptive Statistics, Year 2018 (pre-pandemic)

**Table S2:** Descriptive Statistics, Year 2020 (pandemic)

| Variable | Obs | Weight | Mean | Std. dev. | Min | Max |
| --- | --- | --- | --- | --- | --- | --- |
| MentalHea~ss | 18,127 | 17826.2759 | .0754519 | .2641263 | 0 | 1 |
| lockdown | 18,127 | 17826.2759 | .0225632 | .1485105 | 0 | 1 |
| Hubei_lock | 18,127 | 17826.2759 | .0219223 | .1464341 | 0 | 1 |
| Xinjiang_l~k | 18,127 | 17826.2759 | .0006409 | .0253086 | 0 | 1 |

**Table S3:** Attrition Analysis: The first column represents <u>the basic statistical information of the observations</u> for the whole sample from the raw CFPS dataset. The second column represents the basic statistics information of the observations for the sample after the missing values of the depression symptoms were dropped. The third column represents the basic statistical information of the observations for the sample after the observations that were included in the 2018 CFPS survey but were not included in the 2020 CFPS survey were dropped. The fourth, fifth, and sixth columns represent the comparisons of the statistics indexes between the first and the second column, the first and the third column, and the second and the third column, respectively. This table is used to demonstrate whether the attrition caused by missing observations affects the national representation of the sample used for analysis.

|  | (1) |  | (2) |  | (3) |  | (4) |  | (5) |  | (6) |  |
| --- | --- | --- | --- | --- | --- | --- | --- | --- | --- | --- | --- | --- |
|  |  |  |  |  |  |  | (1) vs (2) |  | (1) vs (3) |  | (2) vs (3) |  |
|  | Mean/<br>SD | Count | Mean/<br>SD | Count | Mean/<br>SD | Count | b/se | *p-value* | b/se | *p-value* | b/se | *p-value* |
| Depression<br>Symptoms<br>(in 2020) | 1.243<br>(1.561)<br>(0.125) | 32598 | 1.243<br>(1.561)<br>(0.124) | 32598 | 1.184<br>(1.505)<br>(0.125) | 19550 | 0.000<br>(0.012)<br>(0.001) | 1.000 | 0.059<br>(0.014)<br>(0.001) | 0.000 | 0.059<br>(0.014)<br>(0.001) | 0.000 |
| Female | 0.500<br>(0.500) | 33097 | 0.500<br>(0.500) | 32598 | 0.501<br>(0.500) | 19550 | 0.000<br>(0.004) | 0.984 | -0.001<br>(0.005) | 0.811 | -0.001<br>(0.005) | 0.798 |
| Age | 44.791<br>(19.366) | 37352 | 44.026<br>(18.588) | 32598 | 42.531<br>(17.745) | 19550 | 0.765<br>(0.144) | 0.000 | 2.260<br>(0.166) | 0.000 | 1.495<br>(0.165) | 0.000 |
| Urban | 0.496<br>(0.500) | 35133 | 0.500<br>(0.500) | 32295 | 0.504<br>(0.500) | 19366 | -0.005<br>(0.004) | 0.242 | -0.008<br>(0.004) | 0.063 | -0.004<br>(0.005) | 0.401 |
| Family Members | 4.170<br>(2.066) | 20924 | 4.179<br>(2.058) | 19895 | 4.180<br>(2.053) | 19529 | -0.009<br>(0.020) | 0.644 | -0.011<br>(0.020) | 0.604 | -0.001<br>(0.021) | 0.954 |
| Education | 1.648<br>(0.677) | 37354 | 1.654<br>(0.669) | 32598 | 1.704<br>(0.672) | 19550 | 0.006<br>(0.005) | 0.231 | -0.044<br>(0.006) | 0.000 | -0.051<br>(0.006) | 0.000 |
| Household<br>Income (~) | 9.634<br>(0.998) | 36683 | 9.672<br>(0.990) | 32266 | 9.705<br>(0.980) | 19425 | -0.037<br>(0.008) | 0.000 | -0.070<br>(0.009) | 0.000 | -0.033<br>(0.009) | 0.000 |
| Marital Status:<br>Never Married | 0.168<br>(0.374) | 19807 | 0.163<br>(0.369) | 18799 | 0.164<br>(0.370) | 18435 | 0.005<br>(0.004) | 0.186 | 0.004<br>(0.004) | 0.340 | -0.001<br>(0.004) | 0.721 |
| Marital Status:<br>Married | 0.770<br>(0.421) | 19807 | 0.776<br>(0.417) | 18799 | 0.776<br>(0.417) | 18435 | -0.006<br>(0.004) | 0.182 | -0.006<br>(0.004) | 0.141 | -0.001<br>(0.004) | 0.887 |
| Marital Status:<br>Cohabiting | 0.003<br>(0.055) | 19807 | 0.003<br>(0.055) | 18799 | 0.003<br>(0.055) | 18435 | -0.000<br>(0.001) | 0.921 | -0.000<br>(0.001) | 0.988 | 0.000<br>(0.001) | 0.934 |
| Marital Status: | 0.018 | 19807 | 0.018 | 18799 | 0.018 | 18435 | -0.000 | 0.987 | 0.000 | 0.939 | 0.000 | 0.927 |
| Divorced | (0.134 |  | (0.134 |  | (0.134 |  | (0.001) |  | (0.001) |  | (0.001) |  |
| Marital Status: | 0.041 | 19807 | 0.040 | 18799 | 0.039 | 18435 | 0.001 | 0.702 | 0.003 | 0.199 | 0.002 | 0.371 |
| Widowed | (0.199) |  | (0.197) |  | (0.193) |  | (0.002) |  | (0.002) |  | (0.002) |  |
| Jobstatus: Agricultural | 0.228 | 37354 | 0.259 | 32598 | 0.260 | 19550 | -0.033 | 0.000 | -0.034 | 0.000 | -0.001 | 0.721 |
| Business | (0.420) |  | (0.438) |  | (0.439) |  | (0.003) |  | (0.004) |  | (0.004) |  |
| Jobstatus: | 0.061 | 37354 | 0.069 | 32598 | 0.071 | 19550 | -0.009 | 0.000 | -0.011 | 0.000 | -0.003 | 0.271 |
| Private Business | (0.239) |  | (0.253) |  | (0.258) |  | (0.002) |  | (0.002) |  | (0.002) |  |
| Jobstatus: | 0.013 | 37354 | 0.014 | 32598 | 0.015 | 19550 | -0.002 | 0.047 | -0.002 | 0.045 | -0.000 | 0.379 |
| Agricultural Job | (0.112) |  | (0.119) |  | (0.120) |  | (0.001) |  | (0.001) |  | (0.001) |  |
| Jobstatus: | 0.245 | 37354 | 0.277 | 32598 | 0.295 | 19550 | -0.034 | 0.000 | -0.052 | 0.000 | -0.018 | 0.000 |
| Wage Job | (0.430) |  | (0.447) |  | (0.456) |  | (0.003) |  | (0.004) |  | (0.004) |  |
| Jobstatus: | 0.010 | 37354 | 0.011 | 32598 | 0.011 | 19550 | -0.001 | 0.129 | -0.001 | 0.348 | 0.000 | 0.722 |
| Unemployed | (0.099) |  | (0.104) |  | (0.102) |  | (0.001) |  | (0.001) |  | (0.001) |  |
| Jobstatus: | 0.119 | 37354 | 0.126 | 32598 | 0.137 | 19550 | -0.008 | 0.001 | -0.019 | 0.000 | -0.011 | 0.000 |
| Student | (0.323) |  | (0.331) |  | (0.344) |  | (0.002) |  | (0.003) |  | (0.003) |  |
| Jobstatus: | 0.092 | 37354 | 0.103 | 32598 | 0.082 | 19550 | -0.012 | 0.000 | -0.009 | 0.000 | 0.021 | 0.000 |
| Retiree | (0.289) |  | (0.304) |  | (0.274) |  | (0.002) |  | (0.002) |  | (0.003) |  |
| Jobstatus: | 0.050 | 37354 | 0.057 | 32598 | 0.058 | 19550 | -0.007 | 0.000 | -0.008 | 0.000 | -0.001 | 0.629 |
| Housework | (0.219) |  | (0.232) |  | (0.234) |  | (0.002) |  | (0.002) |  | (0.002) |  |
| Jobstatus: | 0.025 | 37354 | 0.028 | 32598 | 0.022 | 19550 | -0.003 | 0.017 | 0.003 | 0.020 | 0.006 | 0.000 |
| Disabled | (0.157) |  | (0.165) |  | (0.146) |  | (0.001) |  | (0.001) |  | (0.001) |  |
| Jobstatus: | 0.011 | 37354 | 0.013 | 32598 | 0.011 | 19550 | -0.001 | 0.084 | 0.001 | 0.491 | 0.002 | 0.036 |
| Other | (0.106) |  | (0.112) |  | (0.102) |  | (0.001) |  | (0.001) |  | (0.001) |  |

**Figure S1:**
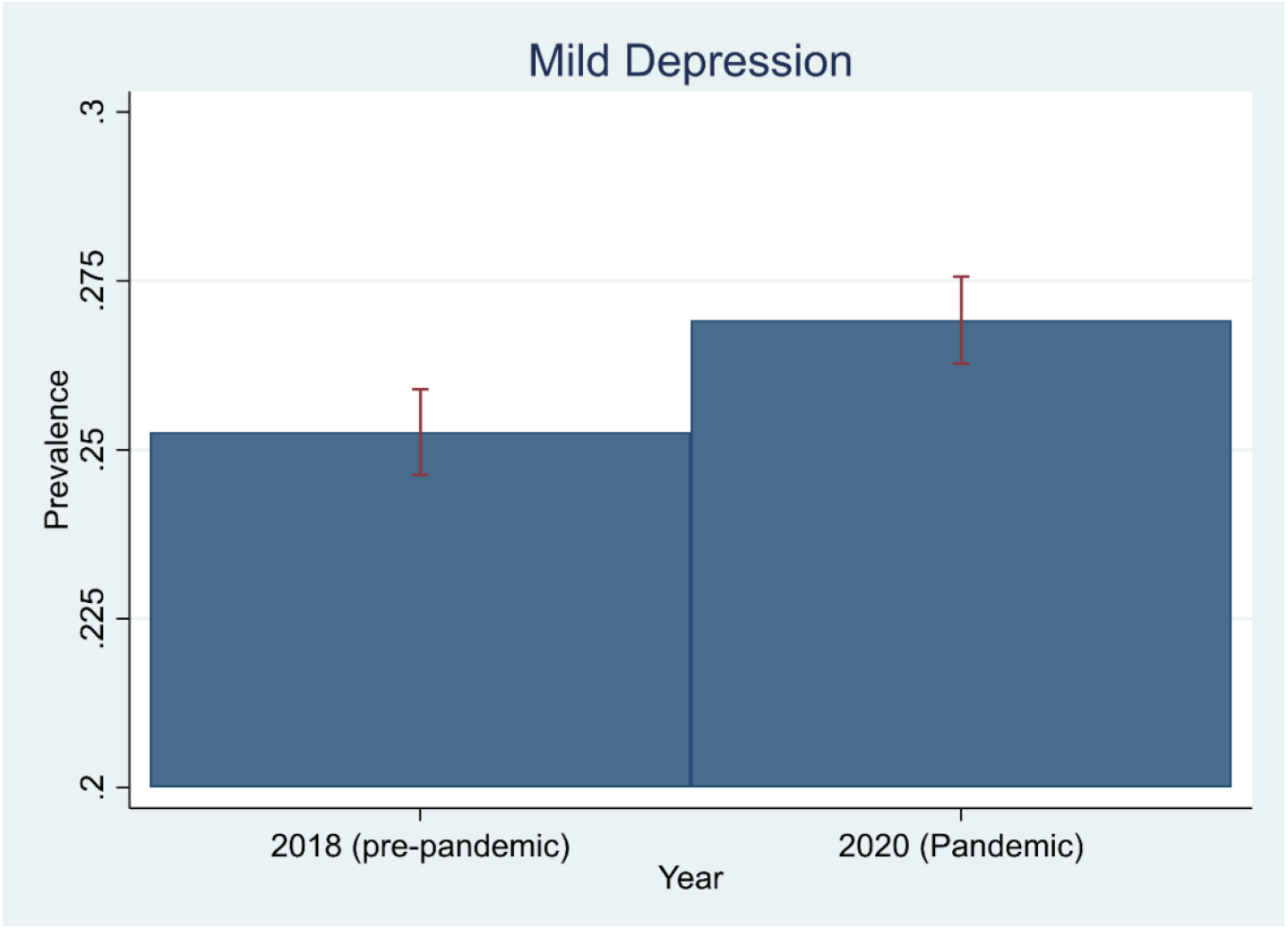
Mild depression in the pre-pandemic and then pandemic waves: Mild depression is defined when responders report 8 or more symptoms in the CES-D questionnaire. The right panel reports the average number of symptoms reported. The bars represent the 95% confidence intervals. Statistics are weighted using the survey sample weights.

**Table S4:** Mental Health and Personality Traits, with reported coefficients on control variables.

| VARIABLES | (1)<br>Personality | (2)<br>+ Basic Demographics | (3)<br>+ Income and Education |
| --- | --- | --- | --- |
| Female |  | -0.060<br>(0.039) | -0.064<br>(0.039) |
| 2.Agegroup |  | -0.030<br>(0.091) | 0.003<br>(0.093) |
| 3.Agegroup |  | -0.096<br>(0.092) | -0.066<br>(0.095) |
| 4.Agegroup |  | -0.050<br>(0.105) | -0.035<br>(0.109) |
| Urban |  |  | 0.036<br>(0.041) |
| size2020 |  |  | -0.000<br>(0.010) |
| 2.Edu_Group |  |  | -0.091*<br>(0.053) |
| 3.Edu_Group |  |  | -0.165* |
|  |  |  | (0.087) |
| logHincome |  |  | -0.028 |
|  |  |  | (0.022) |
| z_Openness | 0.083*** | 0.077*** | 0.081*** |
|  | (0.024) | (0.023) | (0.024) |
| z_Agreeableness | -0.018 | -0.016 | -0.016 |
|  | (0.024) | (0.024) | (0.024) |
| z_Conscientiousness | -0.017 | -0.020 | -0.019 |
|  | (0.027) | (0.027) | (0.027) |
| z_Neuroticism | -0.054** | -0.048** | -0.051** |
|  | (0.022) | (0.023) | (0.023) |
| z_Extraversion | -0.012 | -0.008 | -0.008 |
|  | (0.025) | (0.025) | (0.026) |
| z_Cognitive | 0.002 | -0.012 | 0.033 |
|  | (0.021) | (0.025) | (0.031) |
| 2.Cmonth | -0.255 | -0.242 | -0.263 |
|  | (0.465) | (0.463) | (0.488) |
| 6.Cmonth | -0.103 | -0.077 | -0.045 |
|  | (0.354) | (0.356) | (0.362) |
| 7.Cmonth | 0.195 | 0.220 | 0.233 |
|  | (0.242) | (0.243) | (0.249) |
| 8.Cmonth | 0.242 | 0.267 | 0.278 |
|  | (0.242) | (0.242) | (0.249) |
| 9.Cmonth | 0.277 | 0.307 | 0.337 |
|  | (0.276) | (0.276) | (0.282) |
| 10.Cmonth | 0.401 | 0.418 | 0.456 |
|  | (0.307) | (0.307) | (0.315) |
| 11.Cmonth | 0.115 | 0.136 | 0.132 |
|  | (0.735) | (0.740) | (0.739) |
| 12.Cmonth | 0.202 | 0.176 | 0.243 |
|  | (0.242) | (0.248) | (0.262) |
| Constant | -0.169 | -0.091 | 0.176 |
|  | (0.241) | (0.259) | (0.332) |
| Observations | 13,068 | 13,068 | 13,005 |

Robust standard errors in parentheses
\*\*\* p<0.01, \*\* p<0.05, \* p<0.1

**Table S5:** Mental Health and Some Demographic Characteristics, with reported coefficients on control variables.

|  | (1) | (2) | (3) | (4) | (5) |
| --- | --- | --- | --- | --- | --- |
| VARIABLES | Overall | 2018<br>Symptoms=0 | 2018<br>Symptoms>0 | Overall with Job<br>Class | plus Aggregate<br>Variables |
| Lockdown | 0.306**<br>(0.122) | 0.105<br>(0.132) | 0.548***<br>(0.180) | 0.285**<br>(0.123) | 0.263**<br>(0.124) |
| Female | -0.044<br>(0.037) | 0.001<br>(0.034) | -0.027<br>(0.056) | -0.070*<br>(0.038) | -0.068*<br>(0.038) |
| 2.Agegroup | 0.138*<br>(0.078) | 0.262***<br>(0.068) | 0.229*<br>(0.133) | 0.197**<br>(0.082) | 0.201**<br>(0.082) |
| 3.Agegroup | 0.030<br>(0.087) | 0.175**<br>(0.087) | 0.137<br>(0.138) | 0.077<br>(0.097) | 0.078<br>(0.098) |
| 4.Agegroup | 0.093<br>(0.102) | 0.157<br>(0.100) | 0.137<br>(0.158) | 0.137<br>(0.118) | 0.130<br>(0.118) |
| Urban | 0.005<br>(0.037) | -0.080**<br>(0.038) | -0.010<br>(0.053) | 0.009<br>(0.039) | 0.007<br>(0.039) |
| size2020 | -0.008<br>(0.009) | -0.032***<br>(0.010) | -0.004<br>(0.013) | -0.009<br>(0.010) | -0.011<br>(0.010) |
| 2.Edu_Group | -0.057<br>(0.041) | -0.253***<br>(0.041) | -0.100*<br>(0.060) | -0.063<br>(0.042) | -0.063<br>(0.042) |
| 3.Edu_Group | -0.209**<br>(0.084) | -0.306***<br>(0.063) | -0.343**<br>(0.139) | -0.221***<br>(0.084) | -0.227***<br>(0.083) |
| logHincome | -0.032<br>(0.021) | -0.094***<br>(0.021) | -0.062**<br>(0.030) | -0.026<br>(0.022) | -0.025<br>(0.023) |
| Married | -0.037<br>(0.061) | 0.078<br>(0.063) | -0.025<br>(0.094) | 0.020<br>(0.077) | 0.020<br>(0.077) |
| Cohabitation | 0.219<br>(0.418) | 0.544<br>(0.538) | 0.278<br>(0.553) | 0.393<br>(0.457) | 0.407<br>(0.462) |
| Divorced | 0.299<br>(0.248) | 0.857**<br>(0.430) | 0.225<br>(0.262) | 0.294<br>(0.254) | 0.291<br>(0.253) |
| Widowed | -0.217<br>(0.136) | 0.221<br>(0.138) | -0.175<br>(0.180) | -0.139<br>(0.145) | -0.138<br>(0.144) |
| 2.Cmonth | -0.764*<br>(0.452) | -0.453<br>(0.464) | -0.696<br>(0.564) | -0.746*<br>(0.450) | -0.760*<br>(0.452) |
| 3.Cmonth | -0.160<br>(0.335) | -0.472<br>(0.410) | 0.134<br>(0.376) | -0.117<br>(0.345) | -0.109<br>(0.344) |
| 4.Cmonth | -0.273<br>(0.380) | -0.516<br>(0.472) | 0.173<br>(0.434) | -0.253<br>(0.385) | -0.253<br>(0.386) |
| 5.Cmonth | -0.418<br>(0.298) | -0.719*<br>(0.385) | -0.202<br>(0.314) | -0.416<br>(0.301) | -0.407<br>(0.300) |
| 6.Cmonth | -0.492<br>(0.347) | -0.464<br>(0.390) | -0.149<br>(0.367) | -0.533<br>(0.355) | -0.516<br>(0.353) |
| 7.Cmonth | -0.282<br>(0.257) | -0.423<br>(0.376) | -0.028<br>(0.143) | -0.275<br>(0.263) | -0.271<br>(0.263) |
| 8.Cmonth | -0.225<br>(0.257) | -0.414<br>(0.374) | 0.075<br>(0.139) | -0.214<br>(0.262) | -0.211<br>(0.262) |
| 9.Cmonth | -0.164<br>(0.273) | -0.255<br>(0.397) | 0.030<br>(0.178) | -0.146<br>(0.278) | -0.134<br>(0.280) |
| 10.Cmonth | -0.235<br>(0.268) | -0.459<br>(0.379) | 0.142<br>(0.172) | -0.208<br>(0.272) | -0.209<br>(0.273) |
| 11.Cmonth | -0.197<br>(0.267) | -0.381<br>(0.382) | 0.189<br>(0.177) | -0.179<br>(0.270) | -0.173<br>(0.271) |
| 12.Cmonth | -0.048<br>(0.290) | -0.684*<br>(0.388) | 0.686**<br>(0.284) | -0.022<br>(0.293) | -0.012<br>(0.294) |
| Agricultural<br>Business |  |  |  | 0.025 | 0.030 |
|  |  |  |  | (0.057) | (0.057) |
| Private Business |  |  |  | -0.051 | -0.050 |
|  |  |  |  | (0.087) | (0.087) |
| Agricultural Job |  |  |  | -0.435*** | -0.431*** |
|  |  |  |  | (0.130) | (0.130) |
| Unemployed |  |  |  | -0.098 | -0.120 |
|  |  |  |  | (0.153) | (0.155) |
| Student |  |  |  | 0.108 | 0.110 |
|  |  |  |  | (0.085) | (0.085) |
| Retired |  |  |  | 0.040 | 0.047 |
|  |  |  |  | (0.080) | (0.080) |
| Housework |  |  |  | 0.040 | 0.038 |
|  |  |  |  | (0.086) | (0.086) |
| Disabled |  |  |  | -0.184 | -0.181 |
|  |  |  |  | (0.129) | (0.130) |
| Other |  |  |  | -0.022 | 0.016 |
|  |  |  |  | (0.248) | (0.247) |
| LogGDPpc |  |  |  |  | 0.088 |
|  |  |  |  |  | (0.077) |
| Density |  |  |  |  | 0.024 |
|  |  |  |  |  | (0.020) |
| logFiscalout_pc |  |  |  |  | -0.122 |
|  |  |  |  |  | (0.091) |
| Constant | 0.663* | 2.103*** | 0.032 | 0.511 | -0.504 |
|  | (0.349) | (0.468) | (0.338) | (0.371) | (0.886) |
| Observations | 16,773 | 7,506 | 9,267 | 16,164 | 16,109 |

Robust standard errors in parentheses
\*\*\* p<0.01, \*\* p<0.05, \* p<0.1

## Footnotes

2 To date China counts 5,226 confirmed deaths that can be attributed to COVID, while in the European Union and the United States there are about 1.15 and 1.06 million confirmed deaths respectively (data from www.ourworldindata.org).

3 A comprehensive review of this large literature is beyond the scope of this paper. We refer the reader to [5] for exhaustive meta-analysis and review of this literature and [4] for an illustration of the models linking personality to depression.

4 From the data, we note that while the initial 2018 sample is represented by responders aged between 9 and 103 years old, our final sample after adding data from 2020 is represented by responders aged between 11 and 93 years.

5 Our 8-item scale is derived from the original 20-item scale, which is a commonly used self-rating scale designed to measure depressive symptomatology [41] and is widely considered a validated instrument for screening depression in older adults [41]. The reduced 8-item scale has been shown to be a valid and reliable instrument to detect depression [42].

6 It must be noted however that some non-representative studies on the Chinese population [6] [49] [50] find a higher prevalence of depression and more symptoms of mental health issues in individuals who are older than 60.

7 A finding is consistent with other studies that are based on the Chinese population [6] [7] [25] [49] [50]

